# A composite measure of cerebral small vessel disease predicts cognitive change after stroke

**DOI:** 10.64898/2026.04.23.26351403

**Authors:** Mahir H. Khan, Stuti Chakraborty, Octavio Marin-Pardo, Giuseppe Barisano, Michael R. Borich, James H. Cole, Steven C. Cramer, Emily E. Fokas, Niko H. Fullmer, Leticia Hayes, Hosung Kim, Amisha Kumar, Emily R. Rosario, Heidi M. Schambra, Nicolas Schweighofer, Myriam Taga, Carolee Winstein, Sook-Lei Liew

## Abstract

Post-stroke cognitive recovery is difficult to predict using focal lesion characteristics alone. The brain’s capacity to maintain cognitive function depends also on structural integrity of the whole brain. One way to measure brain health is through the severity of cerebral small vessel disease (CSVD) markers, which reflect aging-related pathologies that erode structural integrity. Here, we propose a composite measure of CSVD (cCSVD) integrating three independently validated biomarkers automatically quantified using T1-weighted MRIs: white matter hyperintensity volume (WMH; representing vascular injury), perivascular space count (PVS; putative glymphatic clearance), and brain-predicted age difference (brain-PAD; structural atrophy). We hypothesize that cCSVD, which captures the shared variance across these CSVD biomarkers, will be a robust indicator of whole-brain structural integrity and predict cognitive changes 3 months after stroke.

We analyzed 65 early subacute stroke survivors with assessments within 21 days (baseline) and at 90 days (follow-up) post-stroke. WMH volume, PVS count, and brain-PAD were quantified from baseline T1-weighted MRIs, and then residualized for age, sex, days since stroke, and intracranial volume. Principal component analysis (PCA) of the residualized biomarkers was used to derive cCSVD. Beta regression with stability selection using LASSO was used to model three outcomes: baseline Montreal Cognitive Assessment (MoCA) scores, follow-up MoCA scores, and longitudinal change (follow-up score adjusted for baseline score). Logistic regression was used to test if baseline cCSVD predicted improvement in those with baseline cognitive impairment (MoCA < 26).

The PCA revealed that the first principal component (PC1) explained 43.1% of the total variance among WMH volume, PVS count, and brain-PAD. The three biomarkers contributed nearly equally to PC1, which was subsequently used as the baseline cCSVD score. Lower baseline cCSVD was significantly associated with better MoCA scores at follow-up (β = −0.19, p = 0.009), even after adjusting for baseline MoCA (β = −0.12, p = 0.042), and, importantly, outperformed all individual biomarkers. Furthermore, lower cCSVD at baseline significantly increased the likelihood of improving to cognitively unimpaired status at three months (OR = 0.34, p = 0.036), independent of age and education.

The composite CSVD captures the additive impact of vascular injury, glymphatic dysfunction, and structural atrophy on recovery in a way that individual measures do not. cCSVD accounts for shared variance across these domains, reflecting a patient’s latent capacity for cognitive recovery, where relative integrity in one CSVD domain may mitigate effects of another. This automated, T1-based framework offers a scalable tool for predicting post-stroke recovery.

## Introduction

Post-stroke cognitive impairment is a primary driver of long-term disability, yet cognitive recovery trajectories are notoriously heterogeneous, making accurate individual prognosis a significant clinical challenge.^1,2^ While demographic factors and focal lesion characteristics are established predictors of initial stroke severity, they do not fully account for the variance associated with long-term cognitive trajectories.^3–5^ This gap has led to growing recognition that the overall health of the whole brain, including structural and vascular integrity, may fundamentally shape the capacity for recovery.^6–8^ Brain health is heavily influenced by natural and pathological aging mechanisms that compromise structural integrity, and has been largely studied by features observable on neuroimaging, such as chronic white matter damage, glymphatic clearance, and global atrophy.^9–11^ While global brain health has been predominantly studied in stroke in relation to motor outcomes or overall functional outcomes (e.g., 90 day modified Rankin Score^12^), these features overlap heavily with those used to characterize cerebral small vessel disease (CSVD), a group of small vessel pathologies that progressively erodes the brain’s structural integrity.^13–15^ Importantly, CSVD markers have been linked to cognitive decline both after stroke and other diseases, such as Alzheimer’s disease.^16–19^ Because cognition is supported by wide-ranging, distributed neural networks,^20,21^ worse brain health especially as measured via CSVD markers likely compromises the brain’s adaptive potential, leading to poorer longitudinal cognitive outcomes after stroke.

On MRI, CSVD is characterized by imaging features including white matter hyperintensities (WMH), enlargement of perivascular spaces (PVS), and brain-predicted age difference (brain-PAD), a measure of brain atrophy.^9,22^ As a marker of chronic white matter damage, WMH represent regions of increased water content, interstitial fluid movement, and, in severe cases, demyelination.^13^ Consequently, larger WMH volume indicates greater CSVD severity and has been associated with worse post-stroke outcomes and poorer response to treatment.^23–25^ Similarly, PVS are fluid-filled compartments surrounding cerebral blood vessels considered to be the main pathway through which glymphatic waste clearance occurs.^26^ While visual rating methods (e.g., counting enlarged spaces on a single representative MRI slice^27^) have historically associated high PVS counts with poor outcomes after stroke,^27^ recent work using automated whole brain PVS segmentations suggests lower counts may indicate existing perivascular occlusion and were correlated with reduced perfusion, marking a loss of structural integrity.^28,29^ Finally, global atrophy resulting from progressive tissue loss can be captured via brain-PAD, a neurobiological construct that compares an individual’s chronological age to age predicted by models trained on structural features from brain MRIs of healthy individuals.^30^ A higher brain-PAD, suggestive of accelerated brain aging, reflects widespread cortical thinning and reduced subcortical volumes, and has been associated with worse post-stroke cognitive outcomes at numerous stages post-stroke.^30–33^

Although mounting evidence links individual CSVD-related biomarkers to post-stroke outcome, they are often examined in isolation.^7,34^ This univariate approach overlooks shared variance across vascular injury, glymphatic clearance, and structural atrophy that together shape the brain’s capacity to respond to injury.^9^ To address this issue, composite measures have been developed to account for varying combinations and severities of CSVD features, recognizing that CSVD is rarely driven by a single biomarker alone.^16,35,36^ However, traditional approaches typically rely on visual rating scales and dichotomized scoring, which can limit sensitivity to the full extent of underlying pathology. Recent improvements in artificial intelligence (AI) have led to more accurate automated methods, which not only save time and reduce the need for researcher expertise, making these measures more accessible, but also provide more sensitive and reliable measurements of CSVD measures, allowing these diverse structural features to be combined into a single, continuous composite score. Such methods may better capture the multidimensional nature of CSVD and enhance prediction of cognitive recovery, which remains difficult to model despite its importance for guiding rehabilitation planning and patient care.^37,38^

In this study, we aimed to generate a composite measure of CSVD (cCSVD) derived using open-source automated AI-driven algorithms, enhancing the accessibility of this technique. We examined whether this cCSVD score, derived via principal components analysis (PCA) of WMH volume, PVS count, and brain-PAD assessed early after stroke, would be associated with cognitive performance at three months post-stroke. We hypothesized that the first principal component would reflect balanced contributions from all three biomarkers, capturing a latent cCSVD construct. We further hypothesized that this composite score would be more strongly associated with cognitive change at three months than any individual biomarker in isolation, providing independent explanatory value beyond initial cognitive performance, demographic information, and infarct volume.

## Materials and Methods

### Participants

Early subacute stroke survivors were recruited from three primary sites: Casa Colina Hospital, Emory University, and NYU Langone Health. Recruitment was based on the following inclusion criteria: (1) presentation with significant upper extremity motor impairment, defined as at least one major muscle group scoring less than 5/5 on the manual muscle test; (2) oral and written proficiency in English or Spanish; and (3) willingness to comply with all study procedures. Individuals were excluded if they presented with traumatic brain injury or other major musculoskeletal or secondary neurological conditions. All study procedures were performed in accordance with the ethical standards of the Declaration of Helsinki, and written informed consent was obtained from all participants prior to enrollment. The Institutional Review Board at the University of Southern California Health Sciences Campus, along with local ethics boards at Casa Colina Hospital, Emory University, and NYU Langone Health, provided ethical approval for this study.

### Data Collection

The study protocol included assessments at two primary timepoints: a baseline assessment conducted within 21 days post-stroke and a follow-up assessment at 90 days post-stroke. While these were the intended milestones, actual timing was adjusted to accommodate participant scheduling. Education level (categorized into three levels: ≤12 years, >12 years, or unknown) and presence of comorbidities such as diabetes, obesity, and hypertension were acquired at baseline. Cognitive performance was evaluated at each timepoint using the Montreal Cognitive Assessment (MoCA). High-resolution T1 structural MRI images were acquired at baseline for all participants using the MPRAGE sequence. Scans were performed on Siemens MAGNETOM Verio and PrismaFit systems with a repetition time of 2300 ms, flip angle of 9°, and variable echo times (2.0, 2.9, or 2.98 ms) and acquisition matrices (224 x 224, 256 x 240, or 256 x 256).

### Image Processing

The baseline T1 images were processed to ensure spatial consistency across subjects. Each T1-weighted image was preprocessed using ANTs’ N4 bias field correction to address intensity non-uniformity,^39^ followed by automated skull stripping with FSL’s BET.^40^ Images were affine registered to the 1 mm isotropic MNI 152 template using skull-stripped masks to constrain registration to brain structures only; this way, extracerebral structures did not influence spatial alignment. Linear affine registration was selected over nonlinear warping to prevent lesion-induced spatial distortions, which can occur when nonlinear algorithms attempt to minimize voxel-wise differences between a lesioned brain and a healthy template.^41^

Following automated processing, all brain masks and registration outputs were manually inspected for accuracy. In cases where automated skull stripping was suboptimal, masks were manually corrected. Furthermore, any scans where the registration quality or brain mask was significantly compromised by the stroke lesion were excluded from subsequent analysis to ensure the integrity of the neuroimaging biomarkers.

Automated cortical reconstruction and volumetric segmentation were performed using the longitudinal FreeSurfer reconstruction pipeline (version 7.4.1).^42^ All segmentations and surface reconstructions were manually inspected for errors, such as inaccurate pial or white matter boundary placement. Any subjects whose reconstructions were significantly distorted by the stroke lesion were excluded from the final analysis.

### Quantification of Neuroimaging Biomarkers

#### Infarct Volume

A trained team of tracers manually segmented stroke infarcts using the T1 image from each visit according to a previously published protocol.^43^ The resulting infarct masks were co-registered to MNI template space using subject-specific linear transformations, and infarct volume was quantified by summing the number of voxels contained within each mask. These volumes were subsequently log-transformed to normalize the distribution.

#### White Matter Hyperintensity Volume

We quantified WMH volume from native-space T1 images using FreeSurfer’s (version 8.0.0) automated WMH-SynthSeg algorithm.^44,45^ WMH-SynthSeg is an unsupervised segmentation method that uses a pretrained convolutional neural network to label brain tissue classes from MRI scans of varying contrast and resolution without retraining, tuning, or preprocessing. The resulting segmentation masks were co-registered to the MNI template space via the previously obtained linear transformations. To ensure that stroke-related tissue damage did not bias the estimates, any voxels within the WMH segmentation that overlapped with the manually traced infarct mask were removed. Following this exclusion, WMH masks were visually inspected to ensure accurate tissue labeling. Total WMH volume was estimated by summing the voxels within the lesion-corrected segmentation mask, then log-transformed to mitigate the right-skew of the volume distribution.

#### Perivascular Space Count

We estimated PVS count using the automated methodology described by Barisano et al.^29^ This approach utilizes a robust segmentation algorithm using native space T1 images as input, leveraging a Frangi-based filter to enhance the tubular structures characteristic of PVS and employing a percentile-based approach to generate PVS binary masks from the vesselness maps. The method identifies and quantifies PVS within the white matter and basal ganglia, providing high accuracy, inter-scanner reproducibility, and test-retest repeatability.^45^ We visually inspected all PVS segmentation masks to verify only PVS structures were captured and minimize the possibility of false positives (e.g., other morphologically similar structures). Subsequent PVS masks were co-registered to the MNI space, and total PVS count was quantified by summing contiguous PVS clusters identified within the white matter and basal ganglia.

#### Brain Age Prediction

Brain age was estimated from the FreeSurfer segmentations using the ENIGMA Brain Age model, which was trained on 4,314 healthy controls and validated in stroke studies.^6,30,46^ This ridge regression model uses 153 regional features including mean cortical thickness, cortical surface area, subcortical volumes, and intracranial volume (ICV) to predict an individual’s brain age. We calculated the brain predicted age difference (brain-PAD) as the difference between the participant’s model-predicted brain age and their chronological age at the baseline assessment. A positive brain-PAD suggests a brain that appears “older” than the participant’s actual chronological age. As quality control (QC), we excluded brain-PAD from any individuals who were missing more than 10% of the FreeSurfer features of interest, based on prior guidance.^6,30^

### Statistical Analysis

#### Principal components analysis

We performed a PCA on WMH volume, PVS count, and brain-PAD from baseline imaging to derive a composite measure of baseline CSVD. Because these three biomarkers are known to vary with demographic factors and head size, each biomarker was first residualized using linear regression with age at baseline, sex, days since stroke, and ICV as covariates. This approach minimized the possibility that any shared variance identified by the PCA was due to these external factors. The resulting residuals were entered into the PCA, and the first principal component was used as the cCSVD score.

#### Feature selection and beta regression modeling of cognitive outcomes

We used beta regression to model MoCA scores at each visit to account for the bounded nature of the measure and its known ceiling and floor effects. To satisfy the (0, 1) interval requirement of the beta distribution, individuals with boundary scores (0 or 30) were excluded, and remaining scores were divided by the maximum score of 30 (hereby referred to as proportional MoCA score).

Three sets of models were estimated. The *baseline outcome model* examined proportional MoCA scores at baseline as the response variable, while the *follow-up outcome model* examined proportional MoCA scores at follow-up. The *change model* examined follow-up proportional MoCA scores while incorporating baseline proportional MoCA scores as a predictor, thereby modeling change in cognitive outcome between visits.^47^ The model equations are as follows:

##### Baseline outcome model

*MoCA(Baseline) ∼ cCSVD(baseline) + age(baseline) + sex*

*+ education + lesion volume + intracranial volume + days since stroke*

*+ diabetes + obesity + hypertension*

##### Follow up outcome model

*MoCA(Follow up) ∼ cCSVD(baseline) + age(baseline)*

*+ sex + education + lesion volume + intracranial volume*

*+ days since stroke + days between scans + diabetes + obesity*

*+ hypertension*

##### Change model

*MoCA(Follow up) ∼ cCSVD(baseline) + MoCA(baseline)*

*+ age(baseline) + sex + education + lesion volume*

*+ intracranial volume + days since stroke + days between scans*

*+ diabetes + obesity + hypertension*

In all models, cCSVD at baseline was included as the primary predictor. We also separately estimated models that included the individual residualized CSVD biomarkers at baseline in place of the cCSVD score. All models were adjusted for covariates previously associated with cognitive performance and post-stroke recovery, including chronological age, sex, education level, lesion volume, and days since stroke.^48^ Additionally, we aimed to incorporate comorbidities based on prior literature evidence of their impact on vascular cognitive impairment.^2,3^ We focused on diabetes, obesity, and hypertension, three primary comorbidities available across the full cohort. Additional analyses related to the impact of comorbidities on cCSVD and cognitive scores can be found in the Supplementary Materials.

However, the inclusion of this extensive set of predictors relative to our sample size raised the significant possibility of overfitting. We thus implemented a stability selection procedure to ensure our findings were both parsimonious and robust.^49^ Proportional MoCA scores were logit-transformed and fit using LASSO (Least Absolute Shrinkage and Selection Operator) regression in the *glmnet* R package, a regularization method that performs variable selection by penalizing the absolute size of coefficients and shrinking non-essential predictors to exactly zero. We executed this procedure over 1,000 random subsamples comprising 80% of the full dataset. Predictors were retained for inclusion in the final beta regression models if they maintained non-zero coefficients in 60% or more of the LASSO iterations. This threshold represents the lower bound of the prescribed stability selection bounds (60% to 90%), balancing predictor sensitivity with the stringency required for high dimensional data.^49^ To maintain a consistent inferential framework, any predictor meeting this threshold in at least one model was retained across all three final models. For categorical variables, the entire factor was included in the final model if any of its constituent dummy-coded levels reached this selection threshold. This approach identified a consistent set of predictors that were then entered into beta regression models to estimate their specific relationships with cognitive outcomes. Prior to analysis, data were assessed to ensure they met the statistical assumptions of beta regression, including the absence of multicollinearity among predictors and the appropriateness of the logit link function. [Line about significance]

#### cCSVD phenotypes and cognitive outcomes

In an exploratory analysis, we investigated whether the phenotypic composition of baseline cCSVD relates to cognitive outcomes. We defined an individual’s cCSVD phenotype based on the directional agreement of the weighted biomarker contributions to their overall cCSVD score. Each biomarker’s contribution was determined as the direction from the product of the standardized residual and its respective PC1 loading. Based on these directions, participants were stratified into three distinct profiles: uniformly low (all biomarkers having negative contributions), uniformly high (all biomarkers having positive contributions), and mixed (divergent directions across biomarkers). We assessed differences in follow-up MoCA scores across these profiles using a Kruskal-Wallis test. This non-parametric approach was selected to account for the relatively small sizes of the exploratory subgroups and the potential non-normal distribution of cognitive scores. Significant global differences were followed by post-hoc Wilcoxon rank-sum tests to evaluate pairwise differences between the phenotypic profiles.

#### Logistic regression for cognitive improvement

We explored the predictive utility of baseline cCSVD by performing a logistic regression focusing on participants with mild cognitive impairment at baseline (MoCA score < 26).^50^ Participants were categorized into two longitudinal trajectories: improved (those scoring ≥ 26 at follow-up) and unchanged (those remaining < 26). The model tested whether cCSVD predicted the probability of improvement while adjusting for age, baseline MoCA, and education level. Education was binarized into two groups based on years of schooling completed (≤12 years vs. >12 years); participants with unknown educational status were excluded to ensure model stability. Model assumptions were verified, showing no evidence of multicollinearity (VIF < 2), no influential outliers (Cook’s distance < 1.0), and confirmed linearity of the logit for all continuous predictors.

## Results

Across all sites, a total of 65 individuals completed both visits and were included in this analysis. This group had a mean (± standard deviation [SD]) age of 58.0 ± 13.7 years at baseline, and 36 individuals (55.4%) were female. Baseline visits occurred at a mean of 24.0 ± 8.9 days after stroke, while follow-up visits took place 96.1 ± 17.6 days after stroke, resulting in a mean interval of 72.0 ± 21.2 days between visits. All imaging data for these individuals underwent rigorous visual QC after each processing step; notably, no individuals were excluded due to failed QC. Infarct volumes in this group were estimated to be 20.3 ± 33.0 mL at baseline. MoCA scores ranged from 0 to 28 at baseline (median: 22, IQR: 8), and 0 to 29 at follow-up (median: 24, IQR: 4.25). Two individuals did not complete MoCA assessments at both visits and were thus excluded from any analyses involving MoCA. These and additional summary statistics are outlined in Supplementary Table 5.

### PC1 reflects balanced contributions from all CSVD biomarkers

We performed a PCA to determine if a cohesive cCSVD score could be derived from the residualized biomarkers. The first principal component (PC1) explained 43.1% of the total variance, while PC2 and PC3 accounted for 29.3% and 27.6%, respectively (Fig. 1A). For PC1, log-transformed WMH volume and brain-PAD loaded together (loadings = 0.55 and 0.61, respectively), whereas PVS count loaded in the opposite direction (loading = −0.57). Importantly, all three biomarkers demonstrated comparable contributions to the PC1 signal (WMH volume: 30.6%; PVS count: 32.5%; brain-PAD: 36.8%), suggesting that PC1 provides a balanced integration of these distinct neuroimaging features.

**Figure 1:**
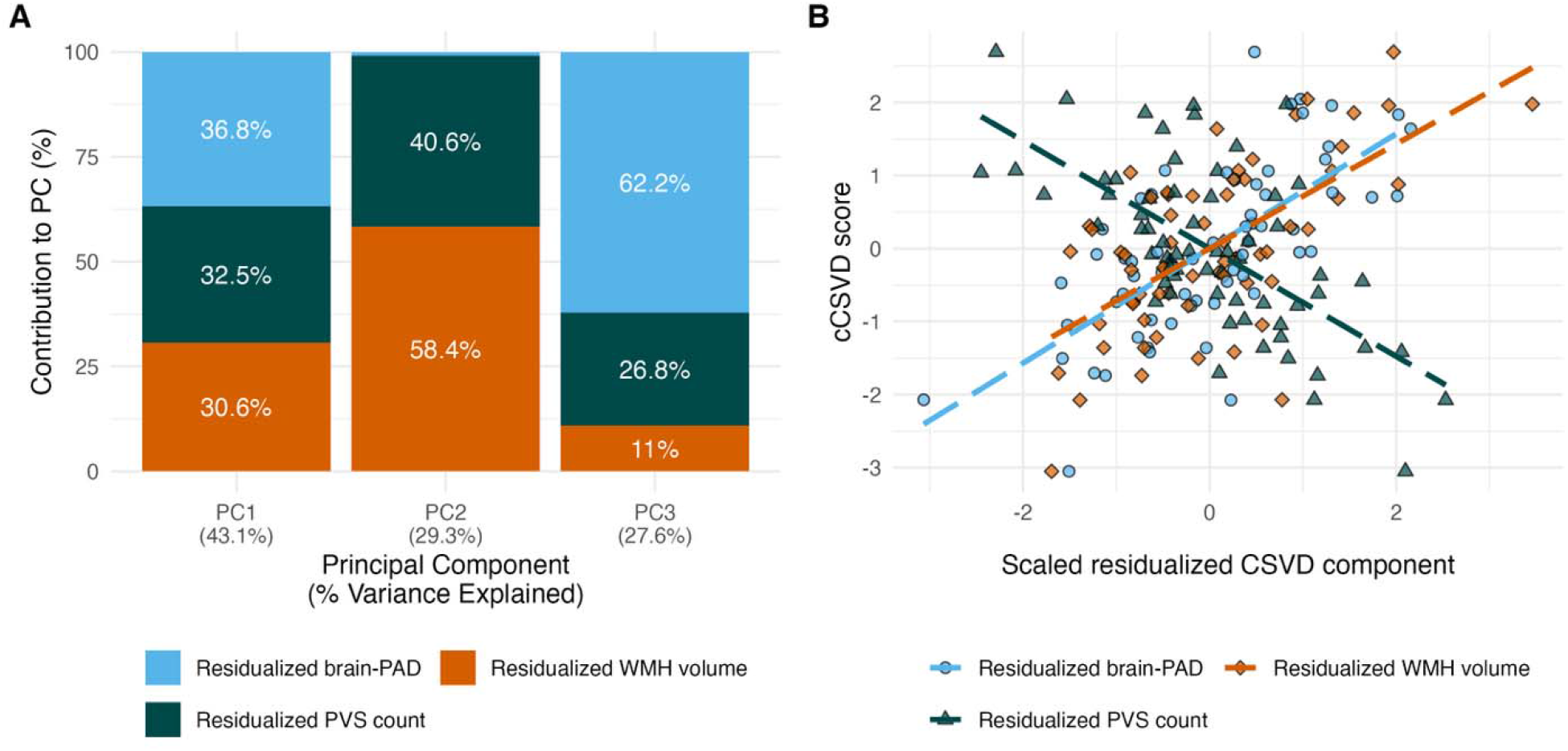
PCA results. (**A**) Contributions of residualized CSVD metrics to each PC. The graph illustrates the relative contribution (%) of residualized WMH volume (orange), PVS count (dark green), and brain-PAD (light blue) to the three principal components. Relative contribution is defined as the squared loading of each variable divided by the sum of squared loadings for that component. The proportion of total variance explained by each PC is indicated in parentheses along the x-axis. PC1 (explaining 43.1% of total variance) shows a relatively balanced contribution from all three metrics, whereas PC2 and PC3 are more heavily weighted toward specific components. (**B**) Correlation of cCSVD score with scaled residualized CSVD components. Individual data points represent participant cCSVD scores plotted against their respective scaled and residualized component values (WMH volume [orange diamonds], PVS count [dark green triangles], and brain-PAD [light blue circles]). Dashed lines indicate the correlation trendline for each component. Higher cCSVD scores correlate with higher WMH volume, higher brain-PAD, and lower PVS count. Abbreviations: brain-PAD: brain predicted age difference; cCSVD: composite cerebral small vessel disease; PC: principal component; PVS: perivascular spaces; WMH: white matter hyperintensities.

Given that PC1 was the dominant individual component and represented a robust consensus across all biomarkers, it was retained as the primary cCSVD metric for subsequent analyses. Within this framework, more positive PC1 scores were characterized by higher WMH volume, higher brain-PAD, and lower PVS counts (Fig. 1B); accordingly, this directional orientation was considered greater CSVD severity.

### cCSVD emerges as a stable predictor after regularized variable selection

Stability selection via LASSO regression identified a robust and parsimonious set of predictors for cognitive function, especially at follow-up (Fig. 2). In the baseline outcome model predicting baseline proportional MoCA using cCSVD as the primary predictor, no predictors reached the 60% threshold for selection; however, cCSVD was the most selected predictor (selection frequency = 50%). For the follow-up outcome and change models predicting follow-up proportional MoCA, cCSVD (≥ 90.3%), age (≥ 86.1%), and education (≥ 74.2%) exceeded the 60% selection threshold. Additionally, in the change model, baseline MoCA was always retained (100%).

**Figure 2:**
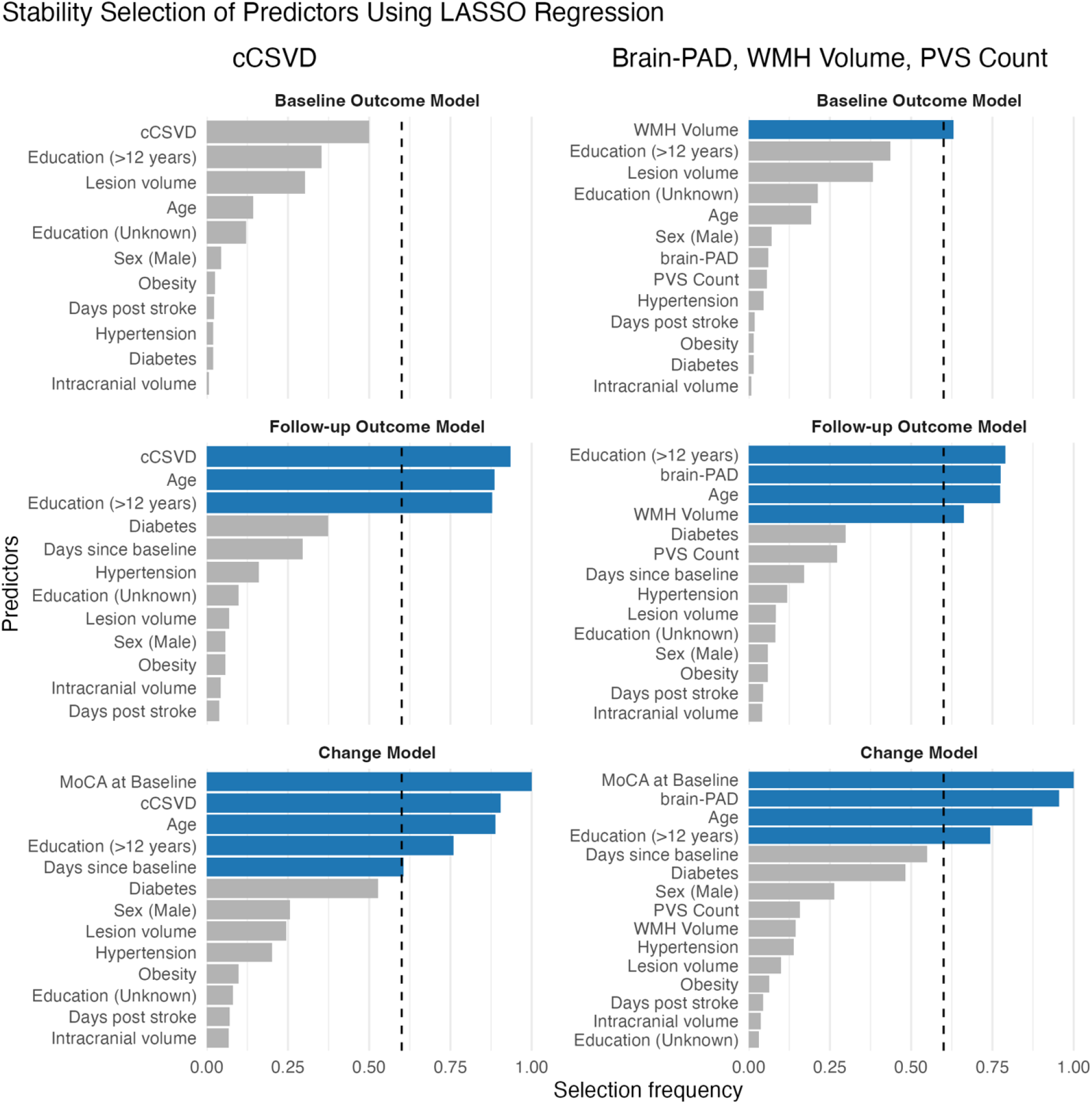
Retained predictors following stability selection using LASSO regression. For each of the three models (baseline outcome, follow-up outcome, and change), LASSO regression was repeated 1000 times using random 80% subsamples of the full dataset. Columns represent stability selection for models on the cCSVD score (left) and the three individual biomarkers (right) as primary predictors. Bars indicate the selection frequency for each predictor across all iterations, with blue bars representing predictors that exceeded the 60% stability threshold (dashed vertical line) for inclusion in final models. For models using cCSVD, no variables achieved the selection threshold in the baseline outcome model; however, cCSVD, age, and education demonstrated high selection stability (>60%) in the follow-up outcome and change models. For the individual biomarker framework, the selected predictors varied across the three models. WMH volume was selected in the baseline outcome and follow-up outcome model, while brain-PAD, age, and education exceeded the selection threshold in the follow-up outcome and change model. MoCA at baseline was also retained in the change model. Abbreviations: brain-PAD: brain predicted age difference; cCSVD: composite cerebral small vessel disease; MoCA: Montreal Cognitive Assessment; PVS: perivascular spaces; WMH: white matter hyperintensities.

When the individual residualized biomarkers were instead used as primary predictors, no single predictor reached the stability threshold across all three models. The WMH volume predictor was selected in the baseline outcome model (63.1%) and follow-up outcome model (66.2%), but not the change model. Meanwhile, brain-PAD, age, and education were retained in the follow-up outcome (77.6%, 77.4%, and 79.0%, respectively) and change (95.5%, 87.2%, and 74.4%, respectively) models. Baseline MoCA was again selected for the change model (100%).

To maintain consistency, a unified set of predictors was applied to all final beta regression models. This set included variables that reached a 60% selection threshold in at least one of the cCSVD or individual component frameworks, resulting in age and education being retained in all models. Although the individual residualized biomarkers (WMH volume, PVS count, and brain-PAD) exhibited differing levels of stability, all three were retained in the individual component models to enable direct comparison with the composite cCSVD results. Lastly, MoCA at baseline was used as a predictor only in the change model.

### Lower baseline cCSVD is associated with higher follow-up cognitive function and change between visits

Following feature selection, baseline cCSVD was significantly associated with cognitive outcomes at follow-up (Table 1; Fig. 3). Although the association with baseline MoCA scores was not significant (*β* = −0.18, *p* = 0.058), higher baseline cCSVD scores were significantly associated with higher follow-up MoCA scores in the follow-up outcome model (*β* = −0.17, *p* = 0.009) and in longitudinal change models adjusting for baseline MoCA (*β* = −0.12, *p* = 0.043). Among covariates, younger age was significantly associated with higher follow up MoCA scores both in the follow up outcome model (*β* = −0.18, *p* = 0.028) and in models adjusting for baseline MoCA (*β* = −0.16, *p* = 0.019).

**Table 1:** Beta regression model summaries for cognitive function using cCSVD following feature selection. Three separate beta regression models are shown for associating proportional MoCA scores at baseline and follow-up with cCSVD (N=62). The three models are: (1) an outcome model predicting MoCA at baseline, (2) an outcome model predicting MoCA at follow-up, and (3) a change model predicting MoCA at follow-up using baseline MoCA as a predictor. To ensure comparability across baseline, follow-up, and change models, a unified set of covariates was used comprising all variables that reached the 60% stability threshold in any of the three outcome-specific LASSO selection procedures. Reported values include beta coefficients, standard errors, 95% confidence intervals, and *p*-values. Lower cCSVD and lower age were significantly associated with higher MoCA at follow-up in both the follow-up outcome model and the change model. Statistically significant coefficients (p < 0.05) are highlighted in bold. Abbreviations: CI = confidence interval, cCSVD: composite cerebral small vessel disease, MoCA: Montreal Cognitive Assessment, SE = standard error.

| Predictor | Baseline Outcome model<br>Proportional MoCA at baseline<br>$n = 62$, Pseudo $R^2 = 0.15$ | | | | Follow-up Outcome Model<br>Proportional MoCA at follow up<br>$n = 62$, Pseudo $R^2 = 0.25$ | | | | Change Model<br>Proportional MoCA at follow-up<br>$n = 62$, Pseudo $R^2 = 0.54$ | | | |
| --- | --- | --- | --- | --- | --- | --- | --- | --- | --- | --- | --- | --- |
| | $\beta$ | SE | 95% CI | $p$ | $\beta$ | SE | 95% CI | $p$ | $\beta$ | SE | 95% CI | $p$ |
| cCSVD at baseline | -0.18 | 0.09 | -0.36 to 0.01 | 0.058 | -0.19 | 0.07 | -0.33 to -0.05 | **0.009** | -0.12 | 0.06 | -0.24 to 0 | **0.043** |
| MoCA at baseline | - | - | - | - | - | - | - | - | 2.07 | 0.32 | 1.45 to 2.7 | **<0.001** |
| Age at baseline | -0.12 | 0.11 | -0.33 to 0.09 | 0.260 | -0.18 | 0.08 | -0.35 to -0.02 | **0.028** | -0.16 | 0.07 | -0.29 to -0.03 | **0.019** |
| Education (>12 years) | 0.31 | 0.23 | -0.14 to 0.76 | 0.178 | 0.33 | 0.18 | -0.02 to 0.67 | 0.066 | 0.21 | 0.14 | -0.07 to 0.5 | 0.137 |
| Education (unknown) | -0.22 | 0.33 | -0.87 to 0.44 | 0.513 | -0.06 | 0.25 | -0.56 to 0.44 | 0.820 | 0.02 | 0.21 | -0.38 to 0.43 | 0.916 |

**Figure 3:**
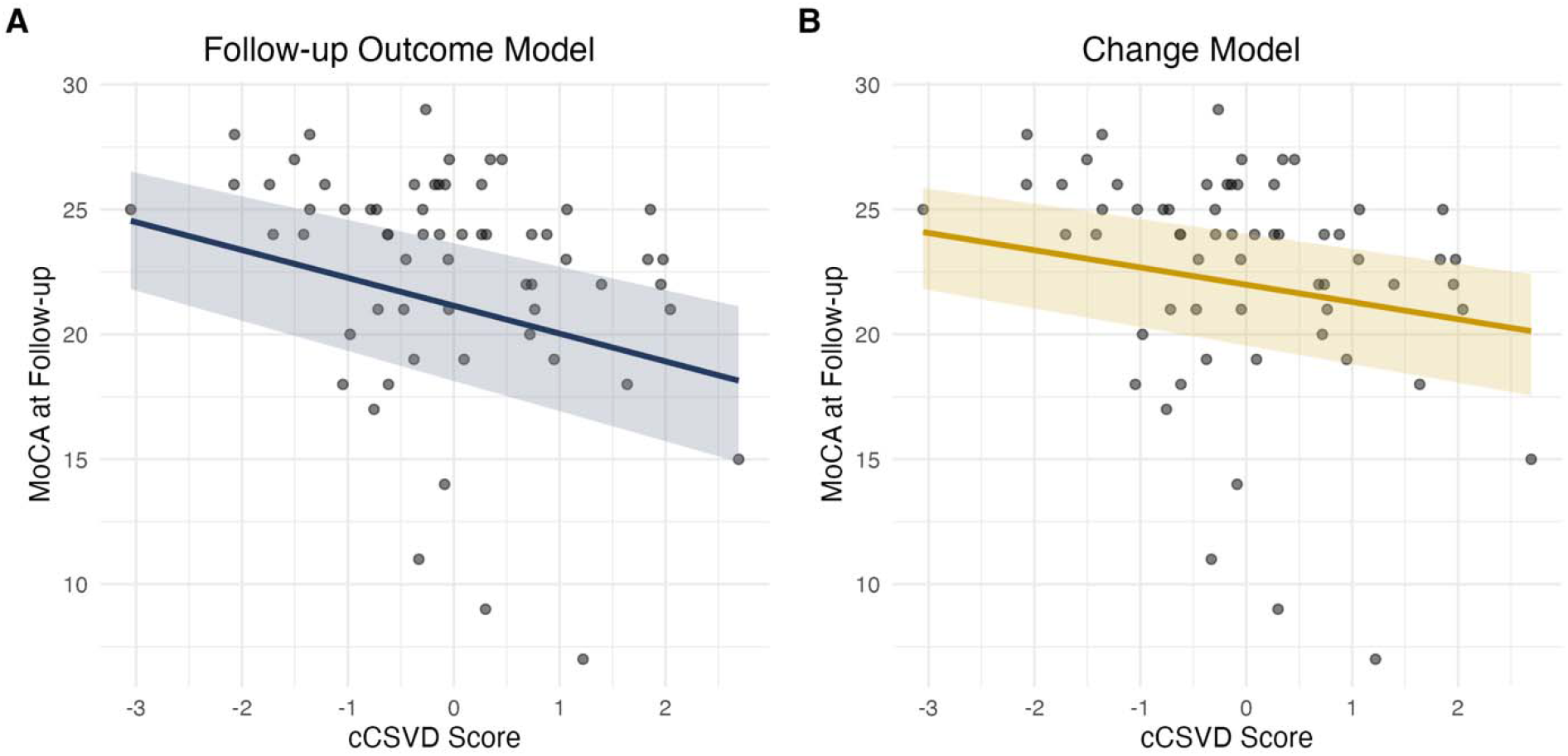
Lower baseline CSVD composite score is associated with better follow-up cognitive function and change between visits. Both panels display individual baseline cCSVD scores plotted against MoCA scores at follow-up (gray points), with solid lines representing predicted mean values from beta regression models and shaded regions representing 95% confidence intervals. (**A**) The follow-up outcome model (navy) associates lower baseline cCSVD scores with higher MoCA scores at follow-up. (**B**) The change model (gold) assesses follow-up MoCA scores conditional on baseline MoCA performance, thereby capturing change between visits. Lower baseline cCSVD scores are associated with higher follow-up MoCA scores after accounting for baseline performance, indicating more favorable cognitive trajectories compared to those with higher scores. Abbreviations: cCSVD: composite cerebral small vessel disease; MoCA: Montreal Cognitive Assessment.

In the models using individual CSVD biomarkers (WMH volume, PVS count, brain-PAD) as predictors, higher WMH volume at baseline was significantly associated with lower baseline MoCA scores (*β* = −0.23, *p* = 0.028), while higher brain-PAD was significantly associated with lower follow-up MoCA scores in the change model (*β* = −0.15, *p* = 0.023; Table 2). No other significant associations were observed between individual CSVD biomarkers at baseline and MoCA at any timepoint. Younger age was significantly associated with higher MoCA scores at follow up (*β* = −0.19, *p* = 0.024), even after accounting for baseline MoCA score (*β* = −0.16, *p* = 0.017).

**Table 2:**
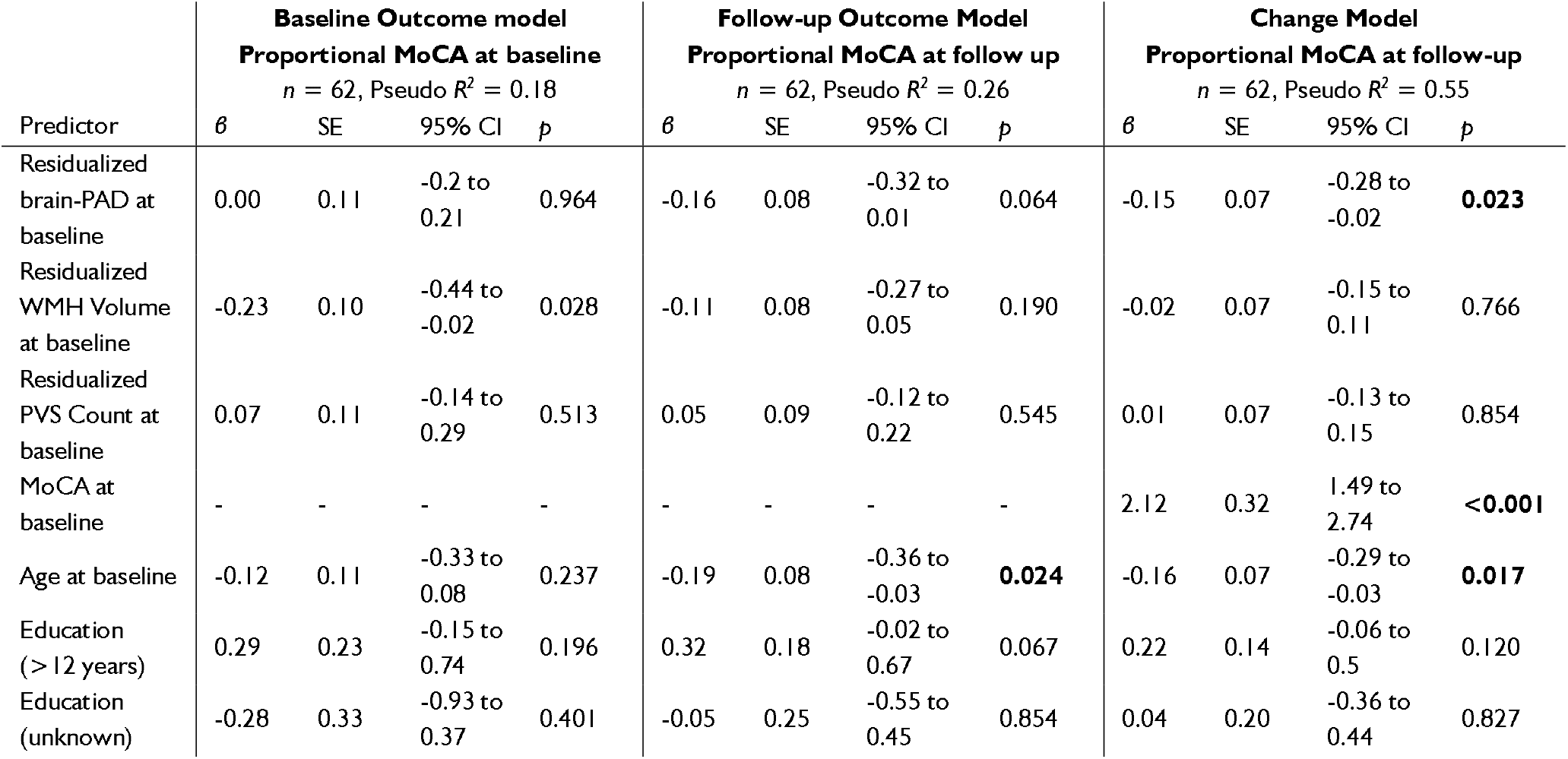
Beta regression model summaries for cognitive function using individual CSVD biomarkers following feature selection. Three separate beta regression models are shown for associating MoCA scores at baseline and follow-up with CSVD biomarkers (N=62). The three models are: (1) an outcome model predicting MoCA at baseline, (2) an outcome model predicting MoCA at follow-up, and (3) a change model predicting MoCA at follow-up using baseline MoCA as a predictor. To ensure comparability across baseline, follow-up, and change models, a unified set of covariates was used comprising all variables that reached the 60% stability threshold in any of the three outcome-specific LASSO selection procedures. Reported values include beta coefficients, standard errors, 95% confidence intervals, and *p*-values. Higher WMH volume at baseline was associated with lower MoCA scores at baseline. Higher brain-PAD at baseline was associated with lower MoCA scores at follow-up in the change model only. Lower age was significantly associated with higher MoCA in the follow-up outcome and change models. Statistically significant coefficients (p < 0.05) are highlighted in bold. Abbreviations: brain-PAD: brain predicted age difference, CI = confidence interval, MoCA: Montreal Cognitive Assessment, PVS: perivascular spaces, SE = standard error, WMH: white matter hyperintensities.

These results were consistent with sensitivity analyses using full models with all candidate predictors included. Full model results are provided in the Supplementary Materials.

### Cognitive outcomes by cCSVD phenotype

In an exploratory analysis of cCSVD phenotypic compositions, an individual’s cCSVD phenotype was defined by the directional agreement of the weighted biomarker contributions to their overall cCSVD score. Each contribution was calculated as the product of the participant’s residualized biomarker score and its respective PC1 loading, with the resulting value indicating a direction toward either higher (positive) or lower (negative) cCSVD severity (Fig. 4A). Participants were then stratified into phenotypes based on the three biomarkers’ directional agreement: those with only negative contributions (uniformly low), those with only positive contributions (uniformly high), and those with divergent contributions across the three features (mixed). A Kruskal-Wallis test revealed significant differences in cognitive outcomes across the three stratified groups (*H*(2) = 10.45, *p* = 0.005). Post-hoc pairwise comparisons using Wilcoxon rank-sum tests demonstrated that the uniformly high group (*n* = 9) had significantly lower MoCA scores than both the mixed (*n* = 44, FDR-corrected *p* = 0.027) and the uniformly low groups (*n* = 12, FDR-corrected *p* = 0.004). Notably, cognitive outcomes for the mixed group did not differ significantly from the uniformly low group (Fig. 4B). A detailed breakdown of biomarker directions in the mixed group phenotype can be found in the Supplementary Materials.

**Figure 4:**
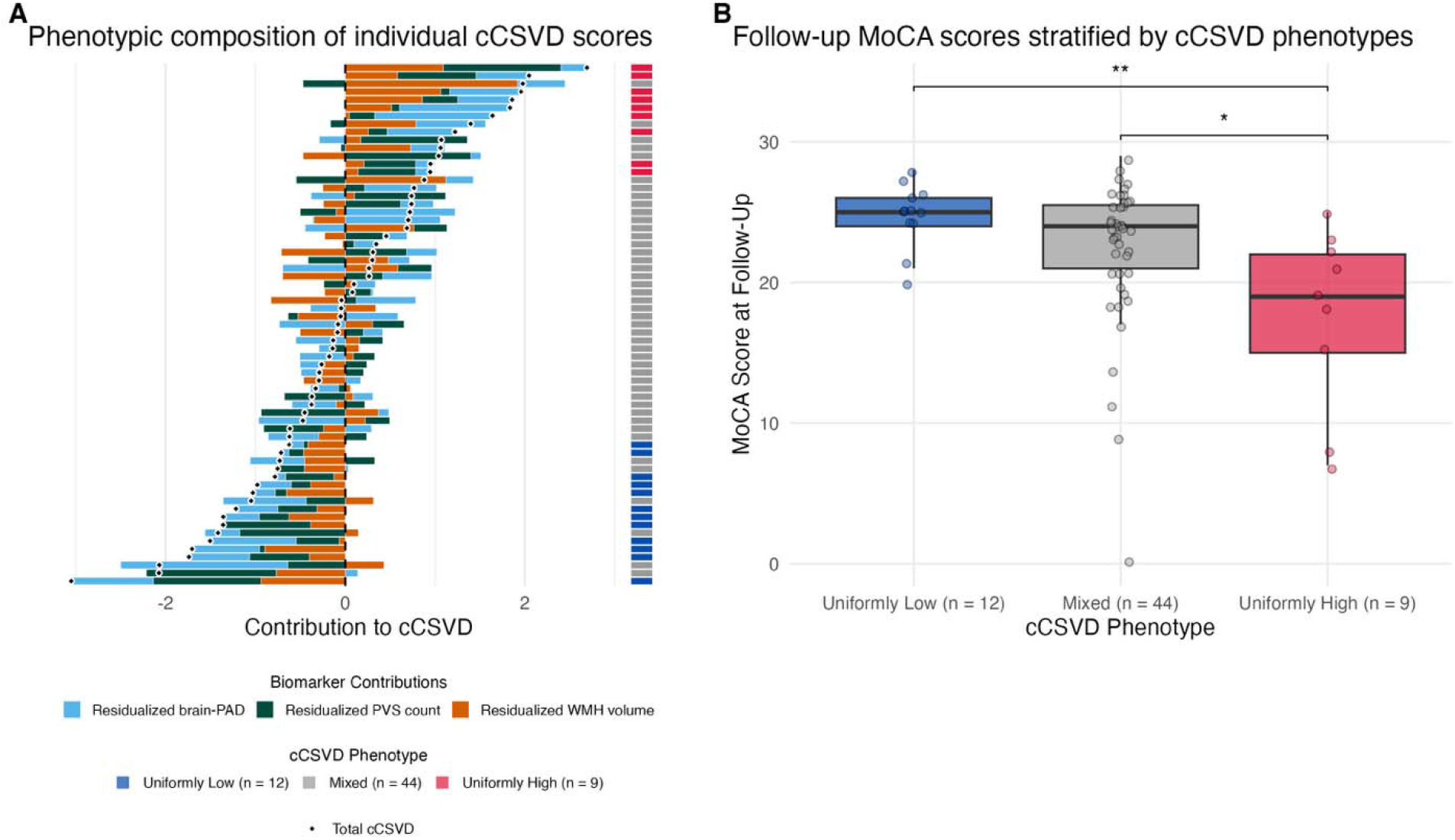
Phenotypic composition of baseline cCSVD scores and its relation to follow-up cognitive function. (**A**) Each horizontal bar represents an individual participant, with the total cCSVD score indicated by the black diamond. The colored segments illustrate the weighted contribution of each neuroimaging biomarker, calculated as the product of the standardized residual value and its respective PC1 loading, to the overall composite score. Bars extending to the left represent negative contributions (indicating lower CSVD severity), while bars extending to the right represent positive contributions (indicating greater CSVD severity). A vertical color strip to the right of the plot indicates each individual’s phenotype based on the directional agreement of their biomarker contributions. Uniformly low (blue, negative) and uniformly high (red, positive) indicate that all three biomarkers contribute in the same direction, while mixed (gray) indicates that at least one biomarker diverges in direction from the others. (**B**) Follow-up cognitive outcomes (MoCA) are stratified by these three cCSVD phenotypes. Box plots demonstrating the median and interquartile range for each group are displayed, with scatterplots indicating each individual’s MoCA score at follow-up. A Kruskal-Wallis test confirmed that cognitive outcomes differed significantly depending on a participant’s cCSVD phenotype. Post-hoc comparisons showed that individuals in the uniformly high group had significantly lower MoCA scores at follow-up than those in either the mixed or uniformly low groups. Notably, there was no statistical difference in performance between the mixed and uniformly low groups.

### Baseline cCSVD predicts improvement to cognitively unimpaired status

Among participants presenting with mild cognitive impairment at baseline (*n* = 47), we examined whether cCSVD predicted improvement to the cognitively unimpaired range (MoCA ≥ 26) at follow-up relative to those who remained unchanged. Logistic regression analysis revealed that lower baseline cCSVD scores significantly increased the likelihood of transitioning from impairment at baseline to the unimpaired range at follow-up (*β* = −1.08, odds ratio = 0.34, *p* = 0.036; Fig. 5). Baseline MoCA, chronological age and education level were not significant predictors of this change (Table 3).

**Table 3:**
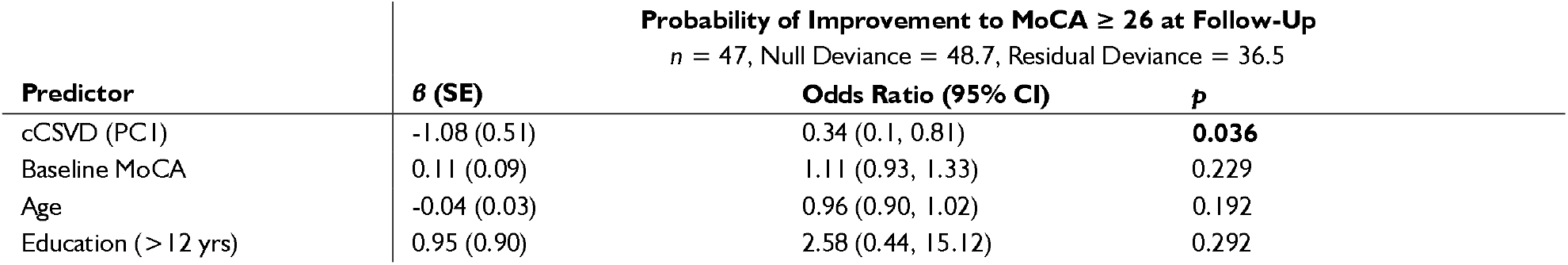
Logistic regression model predicting improvement to cognitively unimpaired status (MoCA ≥ 26) at follow-up. Individuals who were cognitively impaired (MoCA < 26) at baseline were included in this analysis (*n* = 47). Individuals who improved to unimpaired status were compared to those who remained cognitively impaired at follow-up. Model statistics are reported as log-odds (*β*) with standard error (SE), odds ratios (OR) with 95% confidence intervals (CI), and *p*-values. Lower cCSVD at baseline is a significant predictor of improvement, while baseline MoCA, age, and education level were not.

**Figure 5:**
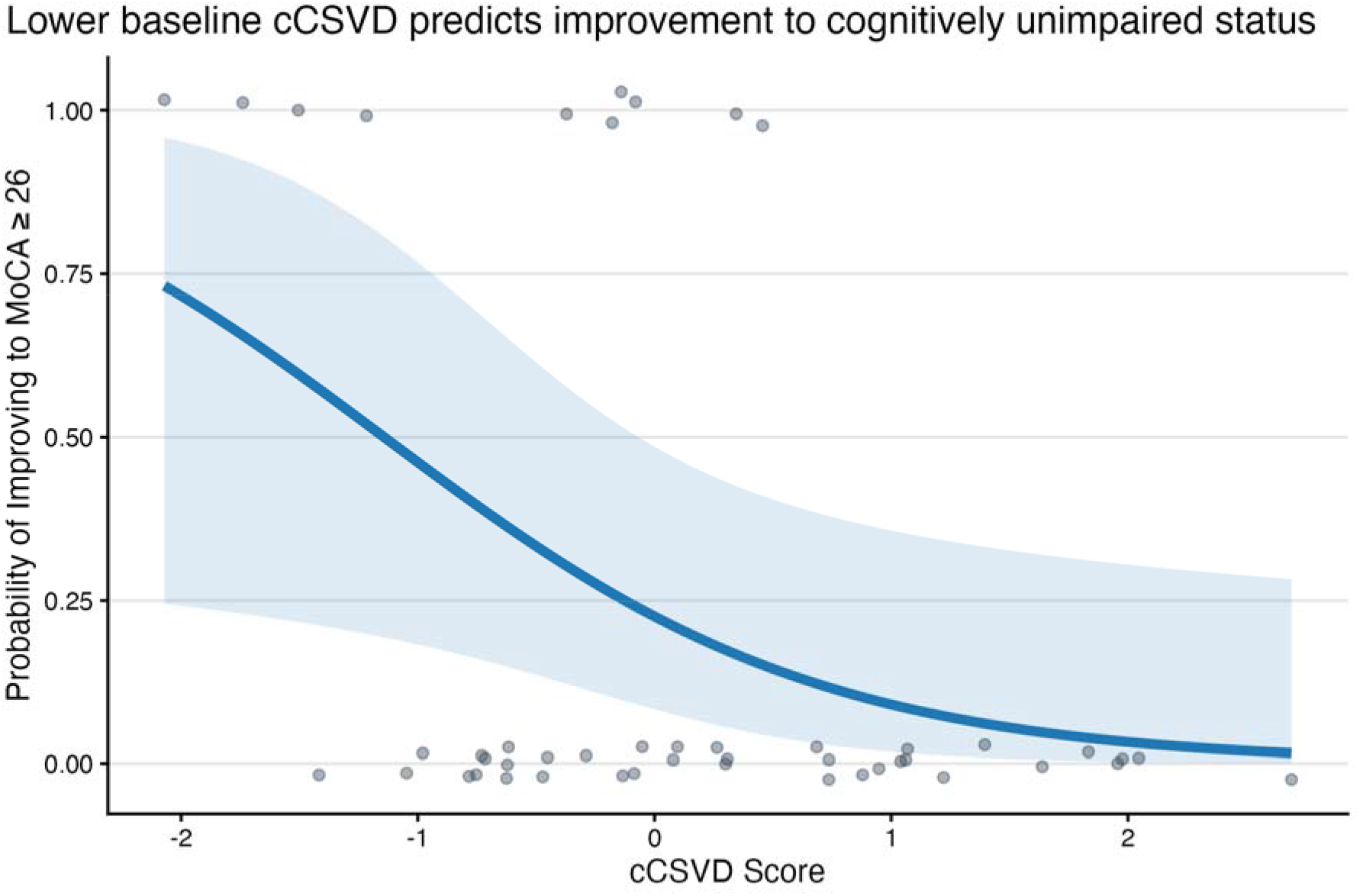
Baseline cCSVD predicts improvement to cognitively unimpaired status. Logistic regression curve illustrating the probability of improving from cognitive impairment (MoCA < 26) to cognitively unimpaired (MoCA ≥ 26) based on cCSVD scores. The blue line represents the predicted probability of improvement across a range of cCSVD scores, with the shaded ribbon indicating the 95% confidence interval. Lower baseline cCSVD scores were significantly associated with an increased likelihood of clinical improvement at follow-up, independent of baseline MoCA, age, and education level. Abbreviations: cCSVD: composite cerebral small vessel disease; MoCA: Montreal Cognitive Assessment.

## Discussion

In this study, we demonstrate that WMH volume, PVS count, and brain-PAD can be combined into a single, cohesive composite score that captures the additive impact of CSVD. Our results suggest that their *collective* signal reflects whole brain integrity and potential for recovery that is a critical determinant of post-stroke cognitive trajectories. The PCA results reveal that these three biomarkers contribute nearly equally to the first principal component, indicating that the estimated cCSVD is not dominated by any single pathological process. Crucially, cCSVD derived shortly after stroke demonstrated a significant relationship with cognitive function and its change to three months post-stroke, even after accounting for baseline status. This association was not consistently observed with each CSVD biomarker individually, indicating that an integrated measure of CSVD may predict an individual’s potential for cognitive change in ways that single imaging biomarkers do not.

Although each biomarker is thought to capture distinct processes related to CSVD, the nearly equal contribution of WMH volume, PVS count, and brain-PAD to the first principal component indicates a high degree of biological alignment among these markers within our study. Rather than any single marker dominating the variance, PC1 was a unified signal where higher brain-PAD and greater WMH volume (representing greater atrophy and accumulated small vessel damage, respectively) loaded in one direction, while higher PVS count loaded in the opposite direction. As noted earlier, although higher PVS count is often clinically assumed to signify poorer brain health, recent automated segmentation literature assessing both larger and smaller MRI-visible PVS proposes that a lower PVS count may represent PVS occlusion and collapse, given that PVS visibility on MRI depends on the presence of fluid in PVS. Lower PVS count is in fact associated with a higher risk of cognitive decline and accelerated brain atrophy in the elderly, and precedes dementia by almost two decades in autosomal dominant Alzheimer’s Disease. Within this framework, a higher count of detectable PVS may reflect a more intact glymphatic clearance system, whereas a reduction in count represents a failure of these channels to effectively manage metabolic waste.^28,29^ As such, the alignment of a lower PVS count with higher brain-PAD and higher WMH volume indicates that the PCA extracted a coherent axis that effectively separates CSVD severity.

The utility of cCSVD is most evident in its relationship with longitudinal cognitive changes following a stroke. We found a significant association between lower cCSVD at baseline and better MoCA performance at three months, even after accounting for baseline MoCA scores, a relationship not observed with any individual biomarker in isolation. Notably, this association remained significant after adjusting for age and education, indicating that cCSVD may capture a unique biological signal for recovery potential beyond traditional demographic proxies. We note that cCSVD did not significantly relate to baseline MoCA scores; baseline cognitive performance may be influenced by several concurrent biological processes, some related to CSVD and some related to acute and subacute stroke-related mechanisms (e.g., delayed necrosis, formation and resolution of edema, neuroinflammatory responses, and diaschisis). At the same time, this also suggests that brain health may not reflect the initial severity of post-stroke cognitive deficits, but rather the brain’s capacity for recovery, which in turn influences the subsequent cognitive trajectory.

Beyond serving as a global marker, cCSVD provides a framework for surveying how specific configurations of biomarker contributions relate to cognitive outcome. Examining these phenotypes reveals that isolated biomarkers can occasionally be misleading; for instance, a single biomarker signaling higher CSVD severity may suggest a high risk of decline that is ultimately mitigated by the relative integrity reflected in other biomarker domains. This was evident in our finding that cognitive outcomes at follow-up were lowest when biomarker contributions were congruent in their positive direction (uniformly high group). In contrast, individuals in the mixed group who had at least one negative contribution demonstrated significantly higher scores, suggesting that even a single CSVD domain reflecting low severity may be sufficient to compensate for higher severity in others. Although this mixed group was highly heterogeneous, representing various combinations of the three contribution directions, the number of individuals within each specific mixed subtype was too small for a meaningful standalone analysis. We were therefore unable to determine if certain direction patterns of the three biomarkers offer unique compensatory advantages. Future work in larger cohorts is needed to confirm whether these distinct phenotypes reflect a true compensatory mechanism, as such research could reveal unique pathways of resilience that are otherwise obscured when evaluating biomarkers in isolation.

In addition to predicting continuous outcomes, our investigation of changes in cognitive impairment status between visits provides further evidence of the prognostic potential of cCSVD. Among participants presenting with baseline cognitive impairment, cCSVD was the sole significant predictor of whether an individual would improve to the cognitively unimpaired range by follow-up. The fact that cCSVD predicted a definitive shift across this established clinical benchmark, especially where demographics and baseline performance did not, is notable. This suggests that CSVD critically affects events underlying stroke recovery, and that therefore early-stage CSVD assessment may offer prognostic utility by identifying individuals with the structural integrity required to reach a clinically recognized stage of recovery.

Another finding of this study is that the relationship between post-stroke cognition and cCSVD appears independent of both demographic factors and comorbidities. To isolate a specific biological signal of structural health, we residualized the individual CSVD biomarkers by age, sex, days post-stroke, and intracranial volume prior to deriving cCSVD. The resulting cCSVD thus captured a state that was not simply a product of demographic or physiological proxies. This was further reinforced by our findings when incorporating comorbidities (see Supplementary Materials); while hypertension status was associated with increases in WMH volume, the presence of comorbidities did not shift cCSVD scores. Furthermore, no clinical risk factors met the threshold for inclusion in our final multivariable models following stability selection. Even when these factors were added into the analysis, the relationship between follow-up MoCA scores and cCSVD persisted, demonstrating that the composite provides further explanatory value beyond a patient’s metabolic or cardiovascular risk status. Collectively, these findings establish cCSVD as an independent and reliable prognostic marker, indicating that the brain’s overall structural state transcends traditional risk profiles in predicting post-stroke cognitive outcomes.

Some limitations of the present study warrant consideration. First, our sample size was sufficient to establish the global utility of cCSVD, but it may have constrained our ability to directly compare the composite against its individual components in multivariable models due to concerns regarding model overparameterization. We sought to mitigate this risk by utilizing a stability selection procedure; notably, even with liberal bounds for variable inclusion, individual biomarkers rarely met the threshold for selection, whereas cCSVD was consistently retained. These results offer preliminary evidence that cCSVD is a more robust predictor in this cohort; however, they should be interpreted with caution until studies with larger cohorts can confirm this finding without constraints on model complexity.

Second, our cCSVD composite was defined by three core imaging markers previously associated with CSVD. While this selection was prioritized to ensure the framework remains scalable and easy to implement across diverse clinical settings, we acknowledge that the full spectrum of small vessel pathology is more expansive. We sought to address this by designing a flexible composite that allows for the seamless integration of additional features, such as lacunes, microbleeds, and cortical superficial siderosis,^13^ as robust automated detection tools become more widely available. These results also establish the broad utility of a whole-brain cCSVD; however, the spatial distribution of the selected markers may be relevant. For instance, WMH or PVS alterations concentrated in strategic white matter tracts may disrupt specific cognitive networks more than a diffuse global load.^9,51,52^ Validation efforts should examine whether incorporating regional brain age predictions or spatial mapping improves predictive specificity.^9,53^

Future work can also evaluate this composite framework against other cognitive assessments. The total MoCA score provides a summary of global cognition that spans several behavioral domains, yet its sensitivity may vary when applied to other global screening tools, to more comprehensive neuropsychological batteries, and to detailed assessments of MoCA’s individual cognitive components (such as memory, visuospatial function, and language).^2,54^ Exploring how cCSVD relates to specific sub-domains, such as executive function or memory,^55^ could reveal how global structural resources interact with localized factors, like specific infarct location, to shape post-stroke outcomes.

Finally, our assessments were limited to three weeks and three months post-stroke. While three weeks is relatively early, imaging may still reflect acute structural changes following the injury, such as resolving edema or rapid secondary atrophy.^56,57^ We attempted to control for this by using days post-stroke as a covariate to residualize our biomarkers. However, future studies should evaluate cCSVD during the hyper-acute phase to determine how the precise timing of assessment influences its prognostic value; doing so would also increase the utility of cCSVD to physicians treating patients during the acute stroke admission. Additionally, post-stroke cognitive trajectories are highly variable; while many patients experience early fluctuations or recovery within the first six months, others face an accelerated risk of future decline or delayed-onset impairment (e.g., vascular dementia) that emerges months or even years after the stroke.^1,58,59^ Given current findings, it is therefore crucial to investigate next whether early-stage cCSVD can identify individuals at risk for these late-onset outcomes, serving as a baseline marker for the brain’s capacity to withstand the progressive neurodegeneration and small vessel disease that drive chronic cognitive decline.

The present work demonstrates that a unified composite measure of CSVD is a more robust predictor of post-stroke cognitive outcomes than its individual biomarker components in isolation. Importantly, these remaining structural resources, when assessed early after stroke, explain significant differences in three-month cognitive impairment and longitudinal change, beyond the influence of demographics, cardiovascular risk factors, and lesion volume. While these findings are focused on subacute MoCA performance, they establish a scalable framework for investigating the role of brain health across broader cognitive domains and more chronic recovery timelines.

## Data Availability

The ENIGMA Brain Age model is available online. WMH-SynthSeg is available as part of the open-source FreeSurfer software package. Additional data and code from this study are available upon reasonable request from the corresponding author.

https://photon-ai.com/enigma_brainage

https://surfer.nmr.mgh.harvard.edu/

## Acknowledgments

During the preparation of this manuscript, the authors used Gemini 3 Pro (Google) to revise the text for clarity and linguistic refinement; the authors reviewed and edited the output as needed and take full responsibility for the final content of the article.

## Supplementary Material

Supplementary materials can be found online.

