## Supplementary Materials for "A composite measure of cerebral small vessel disease predicts cognitive change after stroke"

### Supplementary Methods

#### Sensitivity analysis incorporating comorbidities

We conducted a series of targeted sensitivity analyses to investigate the potential confounding or moderating influence of comorbidities on the relationship between cognitive outcomes and cCSVD. We focused on diabetes, obesity, and hypertension, three primary comorbidities available across the full cohort.

We first compared mean cCSVD scores across comorbidity groups using independent t-tests. To ensure that the composite nature of cCSVD did not obscure individual component effects, we further conducted component-level sensitivity analyses using t-tests to compare individual residualized biomarkers (brain-PAD, WMH volume, and PVS counts) between comorbidity groups. We then evaluated the categorical distribution of cCSVD direction profiles (uniformly unfavorable, mixed, uniformly favorable) relative to comorbidity status. We employed Fisher’s exact tests to evaluate the independence of these profiles from comorbidities, as this method provides robust significance testing for potentially small sample sizes after grouping.

We also examined whether comorbidities moderated any association between MoCA and cCSVD. We implemented Aligned Rank Transform ANOVA models to test for interactions between CSVD direction patterns and each comorbidity on MoCA scores. This non-parametric approach was selected to accommodate the bounded and skewed nature of the MoCA scores while providing a robust framework for interaction effects.

#### Sensitivity Analysis of Recruitment Site Effects

To evaluate the potential impact of recruitment site on our primary findings, we performed a sensitivity analysis comparing our full models with ones that included recruitment site as an additional covariate. We assessed whether the inclusion of site influenced the consistency of the coefficients for cCSVD and the individual biomarkers. These comparisons were used to verify that the observed relationship between brain health and cognitive recovery was driven by a robust biological signal rather than institutional variance.

### Supplementary Results

#### Independence of cognitive outcomes and cCSVD from comorbidities

We evaluated the influence of diabetes, obesity, and hypertension status to determine if comorbidities confounded or moderated the relationship between cCSVD and cognitive outcomes. Statistical comparisons using t-tests showed that mean cCSVD scores were independent of individual comorbidity status. At the component level, sensitivity analyses revealed that greater residualized WMH volume was significantly associated with hypertension (t = -3.27, p = 0.002) and aggregate comorbidities (t = -2.86, p = 0.014); however, no other significant associations were observed between individual comorbidities and residualized CSVD components.

Moving to categorical patterns, Fisher’s exact tests revealed that cCSVD profiles (uniformly unfavorable, mixed, and uniformly favorable) were not significantly associated with the presence of diabetes, obesity, or hypertension. Finally, Aligned Rank Transform ANOVA models indicated that significant group differences in MoCA scores across cCSVD direction patterns remained present (p < 0.05). However, no significant main effects or interactions were observed for any individual comorbidity on follow-up MoCA scores.

#### Site-Invariance of Post-Stroke Cognitive Recovery

To ensure robustness of our findings, we compared our models with all covariates with ones including recruitment site as a covariate (Supplementary Table 1, Supplementary Table 2, Supplementary Table 3, Supplementary Table 4). A comparison of site-inclusive and site-agnostic models revealed that while site was associated with baseline cognitive status, it did not meaningfully contribute to the prediction of longitudinal recovery. Specifically, the inclusion of site in the baseline outcome model using cCSVD as the primary predictor increased the Pseudo R^2^ from 0.21 to 0.33, largely driven by higher baseline MoCA scores at Site 3 (*β* = 0.72, *p* = 0.003). However, in the change model, the inclusion of site failed to improve the model fit. The total variance explained remained at 0.59 for both frameworks, and site was not a significant predictor.

The predictive power of the cCSVD composite remained robust across all frameworks. In the follow-up outcome model, the cCSVD score remained a significant predictor even when accounting for site (*β* = -0.17, *p* = 0.019), whereas individual biomarkers in isolation demonstrated inconsistent stability and lower significance when site was included. Given that site primarily reflected idiosyncratic baseline differences and provided no further explanatory power for the longitudinal recovery process, it was excluded from the primary analyses to prioritize model parsimony.

|  | **Baseline Outcome model**  **Proportional MoCA at baseline**  n = 62, Pseudo R^2^ = 0.21 | | | | **Follow-up Outcome Model**  **Proportional MoCA at follow-up**  n = 62, Pseudo R^2^ = 0.30 | | | | **Change Model**  **Proportional MoCA at baseline**  n = 62, Pseudo R^2^ = 0.59 | | | |
| --- | --- | --- | --- | --- | --- | --- | --- | --- | --- | --- | --- | --- |
| Predictor | β | SE | 95% CI | p | β | SE | 95% CI | p | β | SE | 95% CI | p |
| cCSVD at baseline | -0.18 | 0.09 | -0.36 to 0 | **0.046** | -0.20 | 0.07 | -0.35 to -0.06 | **0.005** | -0.11 | 0.06 | -0.23 to 0 | **0.047** |
| MoCA at baseline | - | - | - | - | - | - | - | - | 2.20 | 0.31 | 1.59 to 2.8 | **<0.001** |
| Diabetes | 0.02 | 0.22 | -0.42 to 0.46 | 0.925 | -0.19 | 0.17 | -0.53 to 0.15 | 0.277 | -0.27 | 0.14 | -0.54 to 0 | **0.048** |
| Obesity | -0.13 | 0.24 | -0.6 to 0.35 | 0.596 | -0.10 | 0.19 | -0.48 to 0.27 | 0.584 | -0.10 | 0.15 | -0.4 to 0.2 | 0.519 |
| Hypertension | 0.12 | 0.27 | -0.41 to 0.66 | 0.650 | 0.25 | 0.21 | -0.16 to 0.66 | 0.227 | 0.22 | 0.16 | -0.1 to 0.54 | 0.172 |
| Lesion volume | -0.22 | 0.11 | -0.43 to 0 | **0.049** | -0.05 | 0.08 | -0.22 to 0.11 | 0.536 | 0.06 | 0.07 | -0.08 to 0.19 | 0.399 |
| Age at baseline | -0.15 | 0.11 | -0.36 to 0.06 | 0.172 | -0.21 | 0.09 | -0.37 to -0.04 | **0.015** | -0.15 | 0.07 | -0.29 to -0.02 | **0.023** |
| Sex (Male) | -0.10 | 0.28 | -0.65 to 0.44 | 0.710 | 0.21 | 0.22 | -0.21 to 0.63 | 0.334 | 0.19 | 0.17 | -0.14 to 0.52 | 0.253 |
| Education  (>12 years) | 0.36 | 0.25 | -0.12 to 0.84 | 0.143 | 0.40 | 0.19 | 0.03 to 0.77 | **0.034** | 0.28 | 0.15 | -0.02 to 0.57 | 0.066 |
| Education (unknown) | -0.05 | 0.37 | -0.77 to 0.66 | 0.883 | 0.12 | 0.28 | -0.43 to 0.67 | 0.662 | 0.21 | 0.22 | -0.22 to 0.64 | 0.339 |
| Intracranial Volume | -0.02 | 0.15 | -0.3 to 0.27 | 0.902 | -0.08 | 0.11 | -0.3 to 0.14 | 0.501 | -0.06 | 0.09 | -0.23 to 0.11 | 0.463 |
| Days since stroke at baseline | 0.04 | 0.10 | -0.16 to 0.24 | 0.718 | 0.11 | 0.10 | -0.09 to 0.3 | 0.287 | 0.08 | 0.08 | -0.07 to 0.23 | 0.285 |
| Days between scans | - | - | - | - | 0.13 | 0.10 | -0.08 to 0.33 | 0.224 | 0.15 | 0.08 | -0.01 to 0.31 | 0.070 |

**Supplementary Table 1: Beta regression model summaries for cognitive function using cCSVD scores and all covariates.** Three separate beta regression models are shown for associating MoCA scores at baseline and follow-up with cCSVD (N=62). The three models are: (1) an outcome model predicting MoCA at baseline, (2) an outcome model predicting MOCA at follow-up, and (3) a change model predicting MoCA at follow-up using baseline MoCA as a predictor. Beta coefficients, standard errors, 95% confidence intervals, and *p*-values are provided for all predictors. Lower baseline cCSVD scores were associated with higher MoCA at baseline and at follow-up, even after accounting for baseline score. Lower lesion volume was associated with higher MoCA scores at baseline, but not at follow-up. Lower age was associated with higher MoCA scores at follow-up, even after accouting for baseline MoCA score. More education was associated with higher scores at follow-up, but not after accounting for baseline MoCA score. Presence of diabetes decreased MoCA scores on average after accounting for baseline score. Statistically significant coefficients (p < 0.05) are highlighted in bold. Abbreviations: CI = confidence interval; CSVD = cerebral small vessel disease; cCSVD = composite CSVD score; MoCA = Montreal Cognitive Assessment; SE = standard error.

|  | **Baseline Outcome model**  **Proportional MoCA at baseline**  n = 62, Pseudo R^2^ = 0.33 | | | | **Follow-up Outcome Model**  **Proportional MoCA at follow-up**  n = 62, Pseudo R^2^ = 0.37 | | | | **Change Model**  **Proportional MoCA at baseline**  n = 62, Pseudo R^2^ = 0.59 | | | |
| --- | --- | --- | --- | --- | --- | --- | --- | --- | --- | --- | --- | --- |
| Predictor | β | SE | 95% CI | p | β | SE | 95% CI | p | β | SE | 95% CI | p |
| cCSVD at baseline | -0.12 | 0.09 | -0.29 to 0.06 | 0.190 | -0.17 | 0.07 | -0.31 to -0.03 | **0.019** | -0.11 | 0.06 | -0.23 to 0 | 0.053 |
| MoCA at baseline | - | - | - | - | - | - | - | - | 2.20 | 0.34 | 1.53 to 2.86 | **<0.001** |
| Diabetes | 0.05 | 0.21 | -0.37 to 0.46 | 0.831 | -0.20 | 0.17 | -0.53 to 0.13 | 0.238 | -0.27 | 0.14 | -0.54 to 0 | **0.046** |
| Obesity | 0.05 | 0.24 | -0.42 to 0.51 | 0.844 | -0.06 | 0.19 | -0.43 to 0.31 | 0.765 | -0.10 | 0.15 | -0.4 to 0.2 | 0.510 |
| Hypertension | 0.17 | 0.26 | -0.34 to 0.67 | 0.522 | 0.30 | 0.20 | -0.1 to 0.7 | 0.139 | 0.22 | 0.16 | -0.1 to 0.54 | 0.178 |
| Lesion volume | -0.13 | 0.11 | -0.36 to 0.09 | 0.255 | -0.01 | 0.09 | -0.19 to 0.17 | 0.921 | 0.05 | 0.07 | -0.09 to 0.2 | 0.473 |
| Age at baseline | -0.21 | 0.10 | -0.41 to 0 | **0.049** | -0.24 | 0.08 | -0.4 to -0.07 | **0.005** | -0.15 | 0.07 | -0.29 to -0.01 | **0.030** |
| Sex (Male) | -0.04 | 0.26 | -0.56 to 0.47 | 0.870 | 0.20 | 0.21 | -0.21 to 0.61 | 0.334 | 0.19 | 0.17 | -0.14 to 0.52 | 0.257 |
| Education  (>12 years) | 0.36 | 0.23 | -0.09 to 0.82 | 0.120 | 0.39 | 0.19 | 0.02 to 0.75 | **0.036** | 0.27 | 0.15 | -0.02 to 0.57 | 0.072 |
| Education (unknown) | -0.12 | 0.35 | -0.8 to 0.57 | 0.741 | 0.08 | 0.27 | -0.45 to 0.62 | 0.763 | 0.21 | 0.22 | -0.23 to 0.64 | 0.345 |
| Intracranial Volume | -0.06 | 0.14 | -0.33 to 0.2 | 0.636 | -0.08 | 0.11 | -0.29 to 0.13 | 0.458 | -0.06 | 0.09 | -0.24 to 0.11 | 0.461 |
| Days since stroke at baseline | -0.04 | 0.10 | -0.24 to 0.15 | 0.649 | 0.11 | 0.10 | -0.08 to 0.3 | 0.245 | 0.08 | 0.08 | -0.07 to 0.23 | 0.292 |
| Days between scans | - | - | - | - | 0.24 | 0.12 | 0 to 0.48 | 0.051 | 0.15 | 0.10 | -0.05 to 0.35 | 0.136 |
| Site 2 | -0.20 | 0.48 | -1.14 to 0.74 | 0.677 | -0.17 | 0.38 | -0.91 to 0.56 | 0.648 | -0.05 | 0.30 | -0.63 to 0.54 | 0.873 |
| Site 3 | 0.72 | 0.24 | 0.24 to 1.19 | **0.003** | 0.42 | 0.22 | -0.01 to 0.85 | 0.057 | -0.01 | 0.20 | -0.39 to 0.37 | 0.955 |

**Supplementary Table 2: Sensitivity analysis of beta regression models for cognitive function including recruitment site.** Three separate beta regression models were estimated to associate MoCA scores at baseline and follow-up with baseline cCSVD (N=62), with the addition of recruitment site as a fixed-effect covariate. The models include: (1) an outcome model predicting MoCA at baseline, (2) an outcome model predicting MoCA at follow-up, and (3) a change model predicting MoCA at follow-up while adjusting for baseline MoCA score. Beta coefficients, standard errors, 95% confidence intervals, and p-values are provided for all predictors. Recruitment site accounted for differences in baseline MoCA scores, but was not associated with differences in follow-up MoCA scores or change between visits. In addition, the predictive value of baseline cCSVD remained robust in the follow-up and longitudinal change models. Consistent with the primary findings, younger age and more education remained associated with higher follow-up scores. Statistically significant coefficients (p < 0.05) are highlighted in bold. Abbreviations: CI = confidence interval; CSVD = cerebral small vessel disease; cCSVD = composite CSVD score; MoCA = Montreal Cognitive Assessment; SE = standard error.

|  | **Baseline Outcome model**  **Proportional MoCA at baseline**  n = 62, Pseudo R^2^ = 0.26 | | | | **Follow-up Outcome Model**  **Proportional MoCA at follow-up**  n = 62, Pseudo R^2^ = 0.31 | | | | **Change Model**  **Proportional MoCA at follow-up**  n = 62, Pseudo R^2^ = 0.60 | | | |
| --- | --- | --- | --- | --- | --- | --- | --- | --- | --- | --- | --- | --- |
| Predictor | β | SE | 95% CI | p | Β | SE | 95% CI | p | β | SE | 95% CI | p |
| Residualized brain-PAD at baseline | -0.07 | 0.11 | -0.28 to 0.15 | 0.550 | -0.15 | 0.09 | -0.33 to 0.02 | 0.083 | -0.11 | 0.07 | -0.24 to 0.03 | 0.116 |
| Residualized WMH Volume at baseline | -0.29 | 0.11 | -0.51 to -0.07 | **0.009** | -0.16 | 0.09 | -0.33 to 0.01 | 0.073 | -0.04 | 0.07 | -0.18 to 0.11 | 0.613 |
| Residualized PVS Count at baseline | -0.03 | 0.12 | -0.26 to 0.2 | 0.795 | 0.04 | 0.09 | -0.14 to 0.22 | 0.681 | 0.04 | 0.07 | -0.1 to 0.19 | 0.559 |
| MoCA at baseline | - | - | - | - | - | - | - | - | 2.21 | 0.31 | 1.59 to 2.82 | **<0.001** |
| Diabetes | 0.04 | 0.22 | -0.4 to 0.47 | 0.864 | -0.18 | 0.17 | -0.52 to 0.16 | 0.305 | -0.26 | 0.14 | -0.53 to 0.01 | 0.058 |
| Obesity | -0.16 | 0.24 | -0.64 to 0.32 | 0.515 | -0.13 | 0.19 | -0.51 to 0.25 | 0.493 | -0.11 | 0.15 | -0.41 to 0.19 | 0.474 |
| Hypertension | 0.35 | 0.29 | -0.22 to 0.92 | 0.225 | 0.32 | 0.23 | -0.12 to 0.77 | 0.151 | 0.21 | 0.18 | -0.14 to 0.56 | 0.238 |
| Lesion volume | -0.26 | 0.12 | -0.5 to -0.02 | **0.030** | -0.08 | 0.09 | -0.26 to 0.1 | 0.360 | 0.04 | 0.07 | -0.1 to 0.19 | 0.553 |
| Age at baseline | -0.18 | 0.11 | -0.39 to 0.03 | 0.092 | -0.22 | 0.09 | -0.39 to -0.05 | **0.010** | -0.15 | 0.07 | -0.29 to -0.02 | **0.024** |
| Sex (Male) | -0.16 | 0.27 | -0.7 to 0.38 | 0.558 | 0.18 | 0.21 | -0.24 to 0.6 | 0.392 | 0.19 | 0.17 | -0.14 to 0.52 | 0.262 |
| Education  (>12 years) | 0.31 | 0.24 | -0.16 to 0.78 | 0.201 | 0.39 | 0.19 | 0.02 to 0.76 | **0.038** | 0.28 | 0.15 | -0.01 to 0.58 | 0.057 |
| Education (unknown) | 0.01 | 0.36 | -0.7 to 0.72 | 0.978 | 0.16 | 0.28 | -0.39 to 0.71 | 0.567 | 0.22 | 0.22 | -0.21 to 0.66 | 0.314 |
| Intracranial volume | -0.02 | 0.14 | -0.3 to 0.26 | 0.901 | -0.08 | 0.11 | -0.29 to 0.14 | 0.501 | -0.06 | 0.09 | -0.23 to 0.11 | 0.481 |
| Days post stroke at baseline | 0.02 | 0.10 | -0.17 to 0.22 | 0.810 | 0.10 | 0.10 | -0.1 to 0.3 | 0.319 | 0.07 | 0.08 | -0.08 to 0.22 | 0.362 |
| Days between scans | - | - | - | - | 0.12 | 0.11 | -0.09 to 0.33 | 0.255 | 0.14 | 0.08 | -0.03 to 0.3 | 0.103 |

**Supplementary Table 3: Beta regression model summaries for cognitive function using individual CSVD biomarkers and all covariates.** Three separate beta regression models are shown for associating MOCA scores at baseline and follow-up with individual CSVD biomarkers (N=62). The three models are: (1) an outcome model predicting MOCA at baseline, (2) an outcome model predicting MOCA at follow-up, and (3) a change model predicting MOCA at follow-up using baseline MoCA as a predictor. Beta coefficients, standard errors, 95% confidence intervals, and *p*-values are provided for all predictors. Lower WMH volume was associated with higher baseline MoCA scores; otherwise no other associations between MoCA scores at any timepoint and individual CSVD biomarkers were observed. Smaller lesion volume was associated with higher MoCA at baseline. Younger age was associated with higher MoCA at follow-up, even after accounting for baseline MoCA. More years of education was associated with higher MoCA at follow-up, but not after accounting for baseline MoCA score. Statistically significant coefficients (p < 0.05) are highlighted in bold. Abbreviations: brain-PAD = brain-predicted age difference; CI = confidence interval; CSVD = cerebral small vessel disease; PVS = perivascular spaces; SE = standard error; WMH = white matter hyperintensities.

|  | **Baseline Outcome model**  **Proportional MoCA at baseline**  n = 62, Pseudo R^2^ = 0.36 | | | | **Follow-up Outcome Model**  **Proportional MoCA at follow-up**  n = 62, Pseudo R^2^ = 0.39 | | | | **Change Model**  **Proportional MoCA at follow-up**  n = 62, Pseudo R^2^ = 0.60 | | | |
| --- | --- | --- | --- | --- | --- | --- | --- | --- | --- | --- | --- | --- |
| Predictor | β | SE | 95% CI | p | Β | SE | 95% CI | p | β | SE | 95% CI | p |
| Residualized brain-PAD at baseline | -0.16 | 0.11 | -0.39 to 0.06 | 0.159 | -0.20 | 0.09 | -0.38 to -0.02 | **0.031** | -0.11 | 0.08 | -0.26 to 0.04 | 0.134 |
| Residualized WMH Volume at baseline | -0.16 | 0.11 | -0.38 to 0.07 | 0.169 | -0.09 | 0.09 | -0.26 to 0.09 | 0.338 | -0.03 | 0.08 | -0.18 to 0.11 | 0.662 |
| Residualized PVS Count at baseline | -0.11 | 0.11 | -0.34 to 0.11 | 0.337 | -0.02 | 0.09 | -0.2 to 0.16 | 0.833 | 0.04 | 0.08 | -0.11 to 0.19 | 0.627 |
| MoCA at baseline | - | - | - | - | - | - | - | - | 2.17 | 0.35 | 1.49 to 2.85 | **<0.001** |
| Diabetes | 0.09 | 0.21 | -0.33 to 0.51 | 0.667 | -0.16 | 0.17 | -0.49 to 0.17 | 0.340 | -0.26 | 0.14 | -0.53 to 0.02 | 0.066 |
| Obesity | -0.03 | 0.24 | -0.49 to 0.44 | 0.907 | -0.09 | 0.19 | -0.46 to 0.28 | 0.623 | -0.11 | 0.16 | -0.41 to 0.2 | 0.491 |
| Hypertension | 0.32 | 0.28 | -0.22 to 0.86 | 0.249 | 0.34 | 0.22 | -0.09 to 0.78 | 0.116 | 0.21 | 0.18 | -0.14 to 0.56 | 0.232 |
| Lesion volume | -0.20 | 0.12 | -0.43 to 0.04 | 0.101 | -0.03 | 0.09 | -0.22 to 0.15 | 0.711 | 0.05 | 0.08 | -0.1 to 0.2 | 0.535 |
| Age at baseline | -0.24 | 0.10 | -0.44 to -0.03 | **0.022** | -0.26 | 0.08 | -0.43 to -0.1 | **0.002** | -0.16 | 0.07 | -0.3 to -0.02 | **0.025** |
| Sex (Male) | -0.08 | 0.26 | -0.59 to 0.43 | 0.765 | 0.19 | 0.21 | -0.21 to 0.6 | 0.353 | 0.19 | 0.17 | -0.14 to 0.52 | 0.260 |
| Education  (>12 years) | 0.37 | 0.23 | -0.08 to 0.82 | 0.109 | 0.42 | 0.18 | 0.05 to 0.78 | **0.024** | 0.29 | 0.15 | -0.01 to 0.59 | 0.059 |
| Education (unknown) | 0.02 | 0.35 | -0.66 to 0.7 | 0.962 | 0.16 | 0.27 | -0.38 to 0.69 | 0.569 | 0.22 | 0.22 | -0.22 to 0.66 | 0.319 |
| Intracranial volume | -0.06 | 0.13 | -0.32 to 0.2 | 0.650 | -0.08 | 0.11 | -0.29 to 0.13 | 0.433 | -0.06 | 0.09 | -0.23 to 0.11 | 0.481 |
| Days post stroke at baseline | -0.07 | 0.10 | -0.26 to 0.12 | 0.482 | 0.09 | 0.10 | -0.1 to 0.28 | 0.373 | 0.07 | 0.08 | -0.08 to 0.22 | 0.367 |
| Days between scans | - | - | - | - | 0.23 | 0.12 | -0.01 to 0.47 | 0.063 | 0.15 | 0.10 | -0.05 to 0.34 | 0.140 |
| Site 2 | 0.01 | 0.49 | -0.96 to 0.97 | 0.991 | 0.02 | 0.39 | -0.74 to 0.79 | 0.955 | 0.01 | 0.31 | -0.6 to 0.63 | 0.962 |
| Site 3 | 0.79 | 0.27 | 0.25 to 1.32 | **0.004** | 0.54 | 0.24 | 0.06 to 1.01 | 0.028 | 0.05 | 0.22 | -0.37 to 0.48 | 0.804 |

**Supplementary Table 4: Sensitivity analysis of beta regression models for individual CSVD biomarkers including recruitment site.** Three separate beta regression models were estimated to associate MoCA scores at baseline and follow-up with individual CSVD biomarkers (N=62), with the addition of recruitment site as a fixed-effect covariate. The models include: (1) an outcome model predicting MoCA at baseline, (2) an outcome model predicting MoCA at follow-up, and (3) a change model predicting MoCA at follow-up while adjusting for baseline MoCA score. Beta coefficients, standard errors, 95% confidence intervals, and p-values are provided for all predictors. Recruitment site accounted for differences in baseline MoCA scores, but was not associated with differences in follow-up MoCA scores or change between visits. Baseline brain-PAD was now associated with follow-up MoCA scores after incorporating site as a covariate, but baseline WMH volume was no longer associated with baseline MoCA scores. Consistent with the site-agnostic findings, younger age and more education remained associated with higher follow-up scores. Statistically significant coefficients (p < 0.05) are highlighted in bold. Abbreviations: brain-PAD = brain-predicted age difference; CI = confidence interval; CSVD = cerebral small vessel disease; PVS = perivascular spaces; SE = standard error; WMH = white matter hyperintensities.

### Additional Supplementary Figures

| **Characteristic** | **Baseline (n=65)** | **Follow-Up (n=65)** |
| --- | --- | --- |
| Age (years) | 58.0 ± 13.7 |  |
| Sex – n (%) |  |  |
| Male | 29 (44.6%) |  |
| Female | 36 (55.4%) |  |
| Education – n (%) |  |  |
| 12 years or fewer | 24 (36.9%) |  |
| Greater than 12 years | 33 (50.8%) |  |
| Unknown | 8 (12.3%) |  |
| Comorbidities – n (%) |  |  |
| Diabetes | 34 (52.3%) |  |
| Obesity | 18 (27.7%) |  |
| Hypertension | 49 (75.3%) |  |
| Days since Stroke | 24.0 ± 8.9 | 96.1 ± 17.6 |
| Days since Baseline Visit |  | 72.0 ± 21.2 |
| Intracranial Volume (cm^3^) | 1439.3 ± 168.2 |  |
| Infarct Volume (cm^3^) – median [IQR] (range) | 6.4 [23.5] (0.2 to 137.2) |  |
| brain-PAD | -3.7 ± 10.5 |  |
| WMH Volume (cm^3^) – median [IQR] (range) | 8.1 [9.0] (1.8 to 106.8) |  |
| PVS Count | 628.6 ± 166.5 |  |
| MoCA – median [IQR] (range) | 22 [8] (0 to 28) | 24 [4.2] (0 to 29) |

**Supplementary Table 5: Summary statistics for demographic, behavioral, and neuroimaging metrics acquired during the study.** All values are reported as mean ± standard deviation (SD) when normally distributed or unless otherwise mentioned. Medians, interquartile ranges [IQR], and ranges are reported for non-normally distributed metrics (Infarct Volume, WMH Volume, and MoCA scores). Demographic characteristics and cardiovascular risk factors were recorded at the baseline visit.

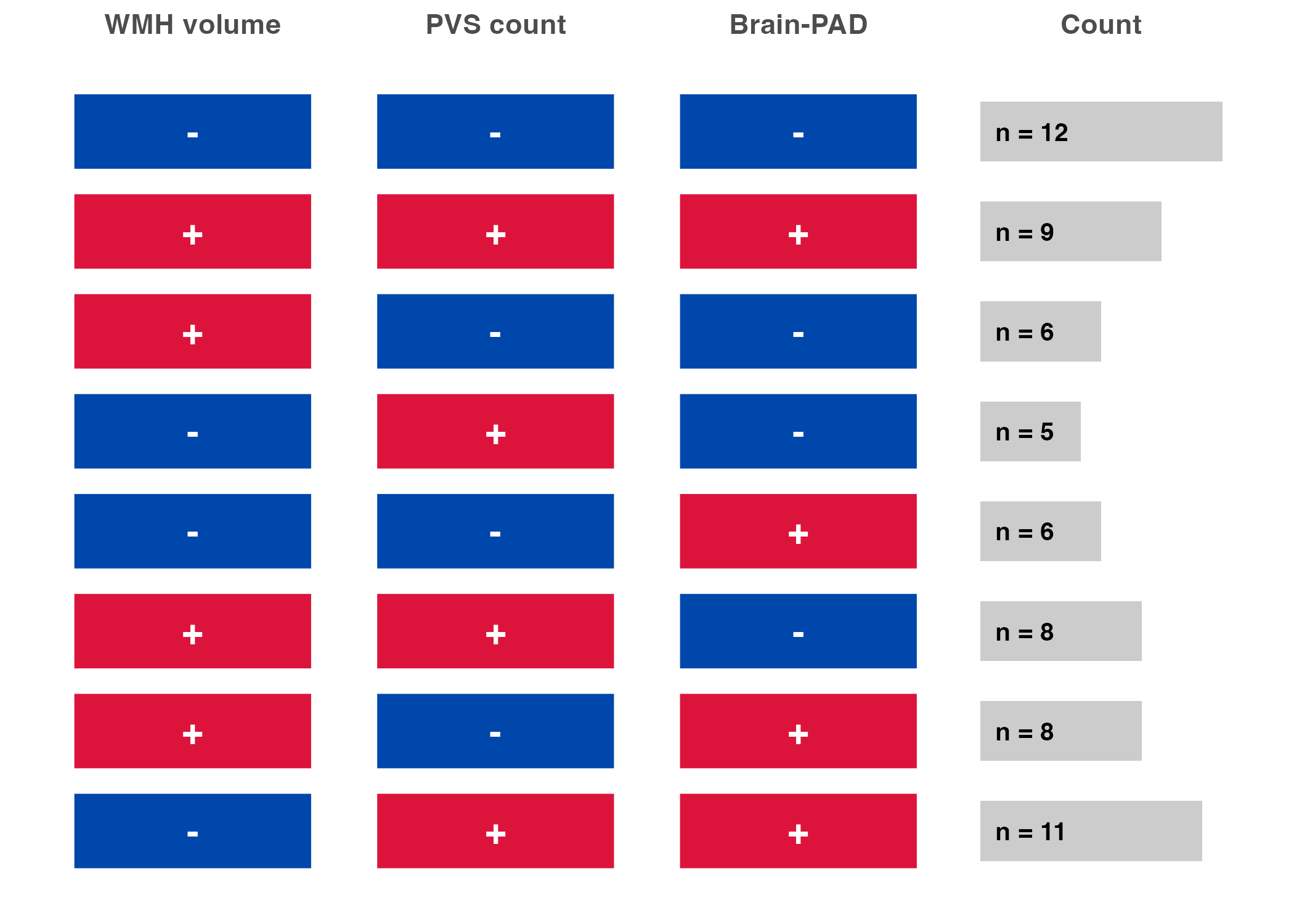

**Supplementary Figure 1: Distribution of cCSVD phenotypes.** This matrix illustrates the eight possible directional combinations of the three underlying biomarkers (WMH volume, PVS count, and brain-PAD) within the study cohort (*n* = 65). Each row represents a unique phenotype, with colors indicating the directional contribution toward the composite cCSVD score (blue [−] = negative direction/lower CSVD severity, red [+] = positive direction/higher CSVD severity). No single phenotype dominates the sample, highlighting the collective CSVD heterogeneity present within the study population.
